# Signal quality and power spectrum analysis of remote ultra long-term subcutaneous EEG

**DOI:** 10.1101/2021.04.15.21255388

**Authors:** Pedro F. Viana, Line S. Remvig, Jonas Duun-Henriksen, Martin Glasstetter, Matthias Dümpelmann, Ewan S. Nurse, Isabel P. Martins, Andreas Schulze-Bonhage, Dean R. Freestone, Benjamin H. Brinkmann, Troels W. Kjaer, Mark P. Richardson

## Abstract

**Objective:** Ultra long-term subcutaneous EEG (sqEEG) monitoring is a new modality with great potential for both health and disease, including epileptic seizure detection and forecasting. However, little is known about the long-term quality and consistency of the sqEEG signal, which is the objective of this study.

**Methods:** The largest multicenter cohort of sqEEG was analyzed, including fourteen patients with epilepsy and twelve healthy subjects, implanted with a sqEEG device (24/7 EEG™ SubQ), and recorded from 23 to 230 days (median 42 days), with a median data capture rate of 75% (17.9 hours/day). Median power spectral density plots of each subject were examined for physiological peaks, including at diurnal and nocturnal periods. Long-term temporal trends in signal impedance and power spectral features were investigated with subject-specific linear regression models and group-level linear mixed effects models.

**Results:** sqEEG spectrograms showed an approximately 1/f power distribution. Diurnal peaks in the alpha range (8-13Hz) and nocturnal peaks in the sigma range (12-16Hz) were seen in the majority of subjects. Signal impedances remained low and frequency band powers were highly stable throughout the recording periods.

**Significance:** The spectral characteristics of minimally-invasive, ultra long-term sqEEG are similar to scalp EEG, while the signal is highly stationary. Our findings reinforce the suitability of this system for chronic implantation on diverse clinical applications, from seizure detection and forecasting to brain-computer interfaces.

**Key Points:**

- Subcutaneous EEG shows similar spectral characteristics to scalp EEG
- The subcutaneous EEG signal is highly stable throughout weeks and months of recording
- Subcutaneous EEG systems are well suited for chronic implantation, for seizure detection and seizure forecasting

## Introduction

Technological advances in recent decades are bringing in a new generation of devices capable of continuous, remote, ultra long-term monitoring of brain activity.^1–4^ The potential applications of these systems are broad, and include monitoring disease activity in dynamic brain conditions such as epilepsy^1,5,6^ or migraine;^7^ brain-computer interfaces for overcoming static neurological deficits in ALS, stroke or spinal cord injury;^8^ or as tools for basic or applied neuroscience research.

Scalp EEG monitoring is limited to a few weeks at most. Electrodes must be held fixed to the skin, with potential for skin injury, inconvenience due to the visibility of the electrode wires, and signal quality degradation with drying of conductive gel and electrode displacement. Mobile EEG caps with dry electrodes^9^ may overcome some of these limitations but signal quality tends to degrade after one hour^.8^ Behind-the-ear or in-the-ear EEG has been studied in patients with epilepsy but its long-term signal quality is unknown^10,11^. All of these systems are also sensitive to physiological artefacts.

Chronic intracranial EEG systems, such as NeuroVista^,1^ RNS NeuroPace^6^ or Medtronic RC+S^4^ have high signal quality with low contribution of physiological artefacts. However, they are invasive systems requiring major procedures and are subject to severe complications, and in some of these^6^ data storage may be limited.

Subcutaneous EEG (sqEEG) systems could offer a trade-off between minimal invasiveness / low risk and good signal quality.^2^ One sqEEG device has been approved in Europe for continuous EEG monitoring, and a single center nine-patient study reported its feasibility and safety in patients with epilepsy.^5^ In a controlled environment, sqEEG signal quality has been shown to be objectively^12^ and subjectively^13^ at least as good as overlapped scalp EEG electrode signals. In addition, several types of artefacts are likely attenuated compared to scalp (sudation, high impedance, some movement artefacts)^.13^ Reliable and stable signal quality is an essential element to the development and validation of automated tools for signal processing, including seizure or spike detection, and seizure prediction^.14–16^ However, the question remains whether signal quality is stable or consistent through time in real-life settings with this system, given the multitude of factors that could deteriorate signal quality.

In parallel, the large amount of subject-specific data generated from ultra long-term sqEEG offers a great opportunity to investigate the long-term intra-individual variability of the human EEG, limited in previous studies by highly-controlled settings and limited data.^17,18^ Stable EEG features would be optimal candidates for disease endophenotypes (“traits”), while highly variable features could be correlates of transient brain “states” or responses to a variety of stimuli.

In this study, analyzing data from a multicenter cohort, we aimed to assess the signal quality and characteristics of an ultra long-term sqEEG system, focusing on signal impedance and power spectral density over time.

## Methods

### Population

Fourteen adult patients diagnosed with refractory epilepsy (median age 44, 66% female) were included from two clinical centres: Zealand University Hospital (ZUH), Denmark (nine patients^5^) and King’s College London (KCL), United Kingdom (five patients from an ongoing seizure forecasting study (ClinicalTrials.gov NCT04061707)). Exclusion criteria were common in both centres: other paroxysmal conditions mimicking epileptic seizures (e.g. psychogenic nonepileptic seizures), lack of evidence for electrographic correlates for seizures, potential clinical risk of device implantation (e.g. concomitant treatment with anti-thrombotic medication), inability to comply with study procedures, low seizure frequency (< 1 seizure/week in the ZUH study and <2 seizures / month in the KCL study) or planned MRI during the study period (for full list of eligibility criteria, see ^5^ and https://clinicaltrials.gov/ct2/show/NCT04061707). One patient from King’s College London (ID S01) underwent reimplantation following a device malfunction, in a different location of implantation to the initial device. We treated this as a separate dataset (ID S01_2). A cohort of twelve adult healthy subjects (Hospital of South West Jutland, DK, median age 35, 58% female) was also included in the analysis. Each study was approved by their institutional review boards, and participants provided written informed consent.

### EEG recording

All subjects used the 24/7 EEG™ SubQ system. The sqEEG implant (UNEEG SubQ™) consists of a 3-contact lead wire (yielding 2-channel bipolar EEG) and a small ceramic housing, implanted unilaterally under local anesthesia, over the region of pre-identified or presumed ictal EEG changes. The three cohorts were inherently unbalanced with regards to location of the implantations (Supplementary Table 1). While different locations were allowed in both patient groups, all healthy control subjects were implanted over the right central region, in a vertical position of the electrode. Recording usually started one to three weeks after implantation. An external recorder connects to the implant housing via an inductive link, powering the implant and storing data. The two-channel EEG signal passes through a uniform amplifier, and the data are recorded with a sampling rate of 207Hz. Impedance measurements are taken during every device connection, usually several times per day. Subjects were asked to use the system as much as possible during their everyday life.

**Table 1.** Differences between healthy subjects and patients with epilepsy, comparing basic demographic features and median frequency band relative powers, after averaging both channels and calculated for the entire 24-hour period.

|  | PWE (n=15*) | HC (n=12) | P value |
| --- | --- | --- | --- |
| Age (median, IQR) | 44 (16) | 35.5 (14.25) | 0.067<sup>+</sup> |
| Gender (Female) | 66% | 58% | 1.000<sup>#</sup> |
| Relative Delta (median, IQR) | 0.45 (0.27) | 0.53 (0.06) | 0.714<sup>+</sup> |
| Relative Theta (median, IQR) | 0.08 (0.04) | 0.11 (0.02) | 0.092<sup>+</sup> |
| Relative Alpha (median, IQR) | 0.08 (0.04) | 0.09 (0.03) | 0.067<sup>+</sup> |
| Relative Sigma (median, IQR) | 0.06 (0.03) | 0.06 (0.02) | 0.575<sup>+</sup> |
| Relative Beta (median, IQR) | 0.21 (0.14) | 0.18 (0.02) | 0.643<sup>+</sup> |

\*One patient underwent two implantations at different sites. These are treated as two separate datasets when comparing relative powers. IQR – Interquartile range; PWE – patients with epilepsy; HC – healthy controls. <sup>+</sup> Wilcoxon rank-sum test. <sup>#</sup> Fisher's exact test.

Median recording duration was 46 days (IQR 41 days), ranging from 23 to 231 days, and was higher in patients than in controls (medians 80 vs. 42 days), due to the different nature of the studies (Supplementary Table 1). Median adherence to the recording device was 75% (average 17.9 recording hours per day), ranging from 25% in one patient and one healthy subject, to 93% in a patient. There was no significant difference in daily adherence between the three cohorts, for the full 24-hour period, (Kruskal-Wallis test, p=0.16), day-time period (p=0.25) and nighttime period (p=0.18), although some subjects showed a predisposition to record more either during the day (e.g. HC04) or during the night (e.g. S04).

### EEG preprocessing and power spectra computation

The EEG was bandpass filtered at 0.5 - 48Hz with a finite-impulse-response equiripple design and 40 dB attenuation filter. The EEG data for each subject were partitioned into non-overlapping 1-minute-long segments. We considered this length as a good balance between temporal resolution and minimizing the effect of random fluctuations in the signal.

The power spectra for each channel and segment were estimated via an adapted version of Welch’s method. Non-overlapping epochs of 4-second duration were mean normalized and tapered with a Hann window. Fast Fourier Transform (FFT) was performed and the frequency range of interest was selected at 0.5 - 40Hz. The magnitude of the FFT coefficients was squared, multiplied by two (for a one-sided spectrum) and scaled by window energy. The power spectral density (PSD) was averaged for every minute segment by taking the median instead of the mean (to reduce the effect of outliers in a highly skewed distribution). Epochs containing flat signal (due to temporary disconnections) or artefacts due to impedance measurements (yielding a signal with high power at 34.5Hz), as well as their adjacent epochs, were excluded from the analysis. Power was converted to dB/Hz units (10*log^10^(power)).

Median power spectra were plotted for each subject-channel pair, over the whole dataset, as well as only for presumed daytime periods (10am to 4pm) and nighttime periods (11pm to 5am), to assess well known power spectral differences between these periods. The power spectrum plots were visually inspected for physiological peaks. In addition, for each segment, we calculated the median absolute power in each of the IFCN-defined (2017 version^19^) canonical frequency bands (delta [0.5-4Hz], theta [4-8Hz], alpha [8-13Hz], beta [13-30Hz]), as well as the sigma band [12-16Hz].

To explore between-group comparisons (patients vs. healthy subjects), frequency band power was converted to relative power (by dividing by the total broadband power of the EEG signal), to account for differences in signal attenuation/volume conduction of different subjects (e.g. different skull thickness or depth of the implant in the subgaleal space), and was averaged between channels.

### Statistical analysis

Descriptive analysis was summarized using median and interquartile ranges (IQR) for continuous variables and counts (percentage) for categorical variables. Between-group differences in relative powers and in demographic information were tested using the Wilcoxon rank-sum test or Fisher’s exact test.

To investigate the long-term stability of sqEEG signal impedance and frequency band absolute power, at the individual level, linear regression models were performed for each subject-channel-signal measure. All impedance values or the daily median of each power measure were set as the dependent variable, and time (in days since start of the recording) as the independent variable. The slope value and the R^2^ of each model were inspected to assess the gradient of change and the fit of the model, respectively.

At the group level, a multilevel linear mixed-effects model was applied. The input variable (and fixed effect to be tested) was time (in days since start of recording), and grouping variables were study subject and sqEEG channel nested within subject. The analysis was repeated splitting daytime and nighttime data, given the different proportions in adherence to these periods by the study subjects and the difference in spectral characteristics. Finally, we explored an effect of patient status (vs. healthy subject) in the stability of the signal by adding a “patient X time” interaction term. All data and statistical analyses were performed in MATLAB (Mathworks, R2020a, USA).

## Results

After exclusion of epochs containing undesired data (as described above and comprising 0.16 to 2.29% of the datasets per patient), a total of 1249 days (41 months) of data from 27 subjects was included in the analysis.

### Power spectral density

Figure 1 shows the median power spectra for each patient from diurnal and nocturnal periods, respectively. Overall, power decreased with increasing frequency, and resembled a 1/f distribution in most study subjects. This was less evident during the daytime period when high frequency power was of larger amplitude, and the 1/f slope flattened (particularly in the beta band). Overall higher broadband power was seen during daytime compared to nighttime. A bi-modal distribution was seen in most patients when plotting the histogram of frequency band power, with each peak being mostly represented by diurnal and nocturnal periods, respectively (Supplementary Figure 1). Even with long-term averaging, peaks in the alpha range during daytime periods were seen in the majority (63%) of subjects (17/27), while peaks in the lower beta / sigma band were seen at nighttime in 16/27 (59%) subjects.

**Figure 1.**
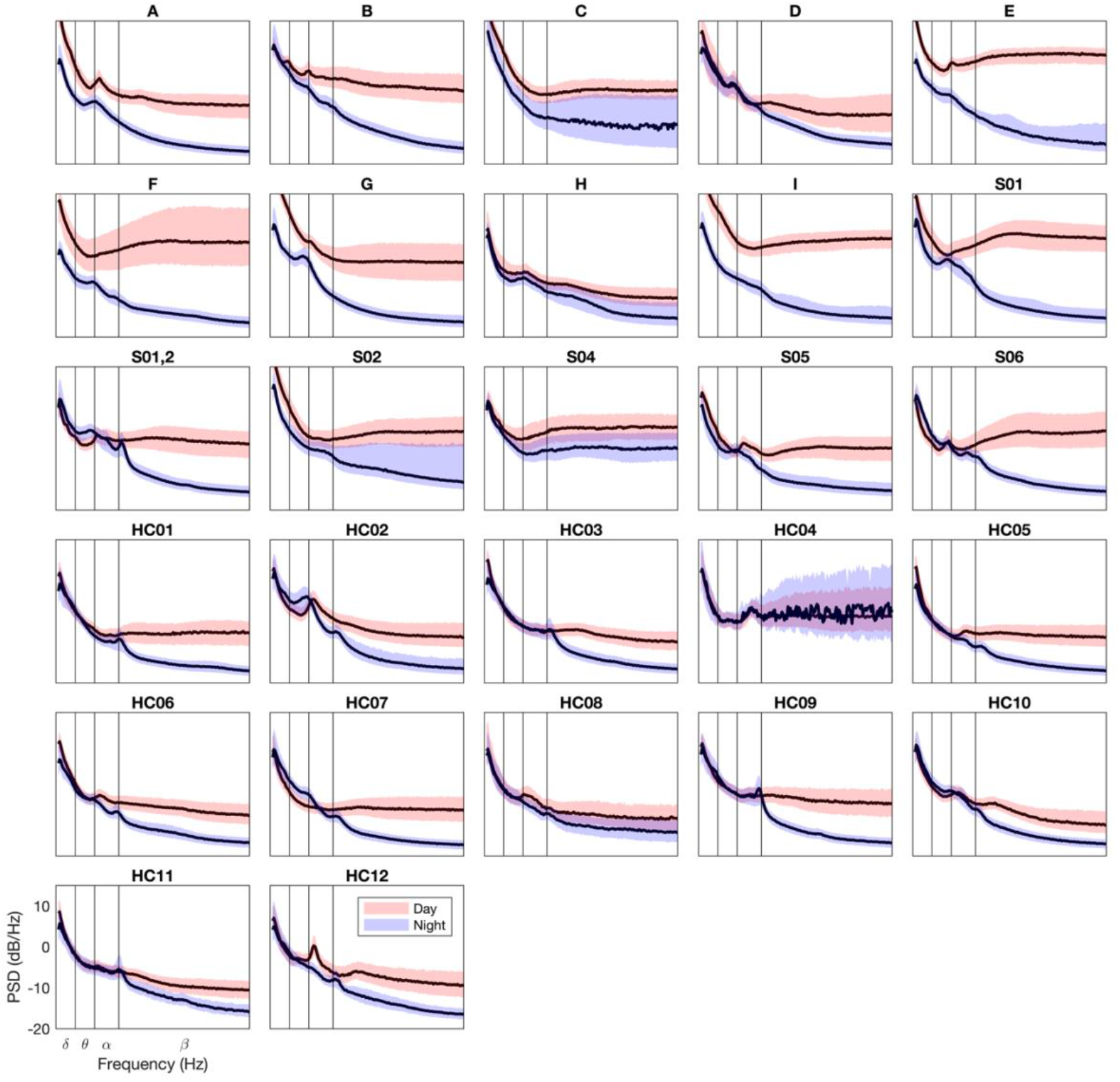
Power spectral density plots for each subject (average of both channels), divided into daytime (10am to 4pm - light red) and nighttime periods (11pm to 5am - light blue). Dark lines represent medians and shaded areas interquartile ranges of the 1-minute segments. Vertical lines represent boundaries of canonical frequency bands. Note that subject H04 recorded little sqEEG during the night and hence data dispersion is large.

### Consistency of the sqEEG signal

The long-term behavior of median daily frequency-band absolute powers was stable for most subjects, albeit some interindividual variability (Figure 2). Individual subject-channel-band linear regression models showed overall gradients close to zero and R^2^ values with low fit (Supplementary Figures 2 and 3). In some subjects there was a slow positive trend (C, I). In others there appeared to be a small increase in the first days, followed by stabilization (F, E, S01).

**Figure 2.**
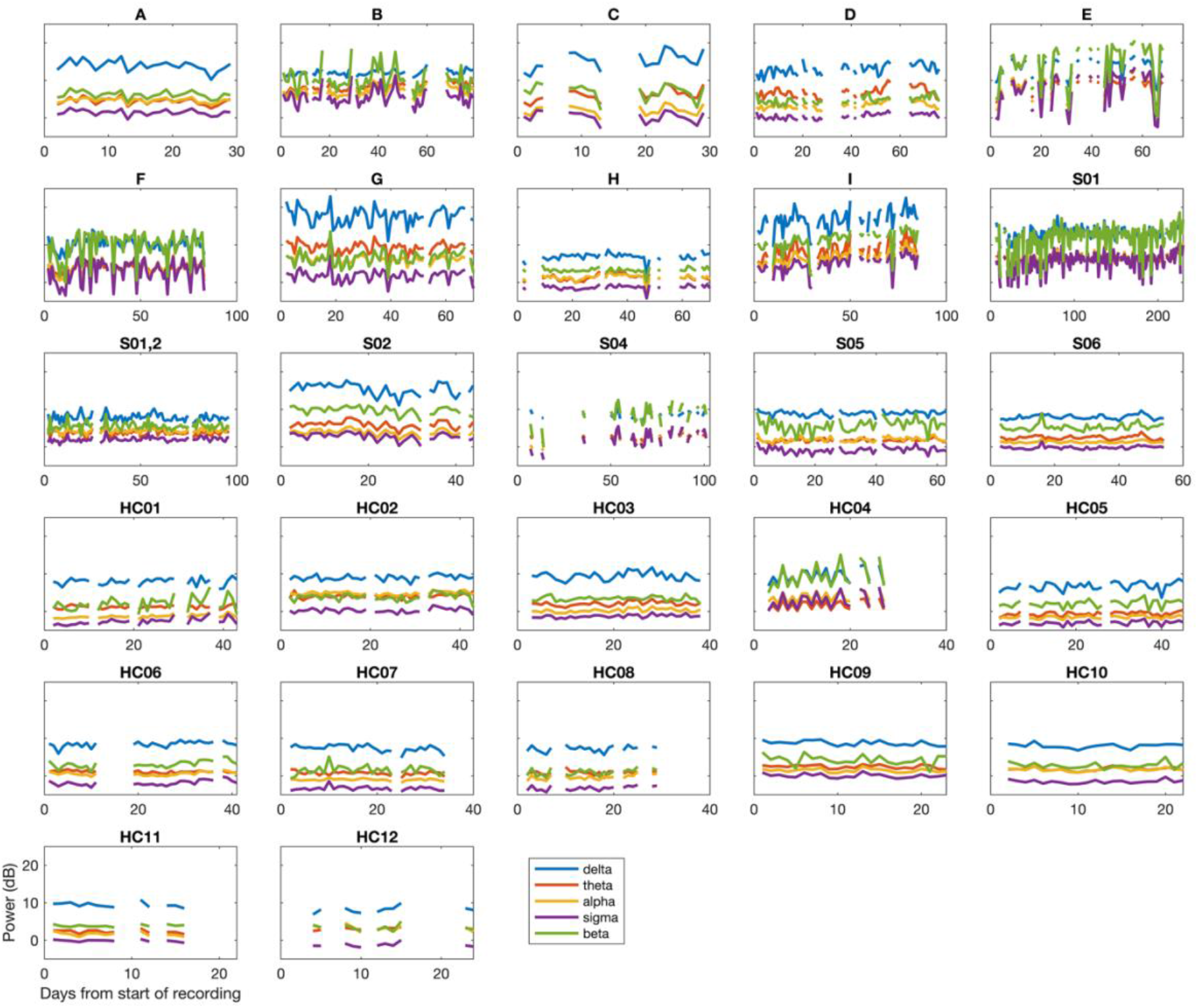
Long-term variation in absolute power (in dB) in different frequency bands. Each plot depicts the median daily power at each frequency band, averaged over both channels, throughout the study period for each subject. Days with low (<50%) adherence were removed from the plots. Note that the study duration (x-axis) is different for each subject.

**Figure 3.**
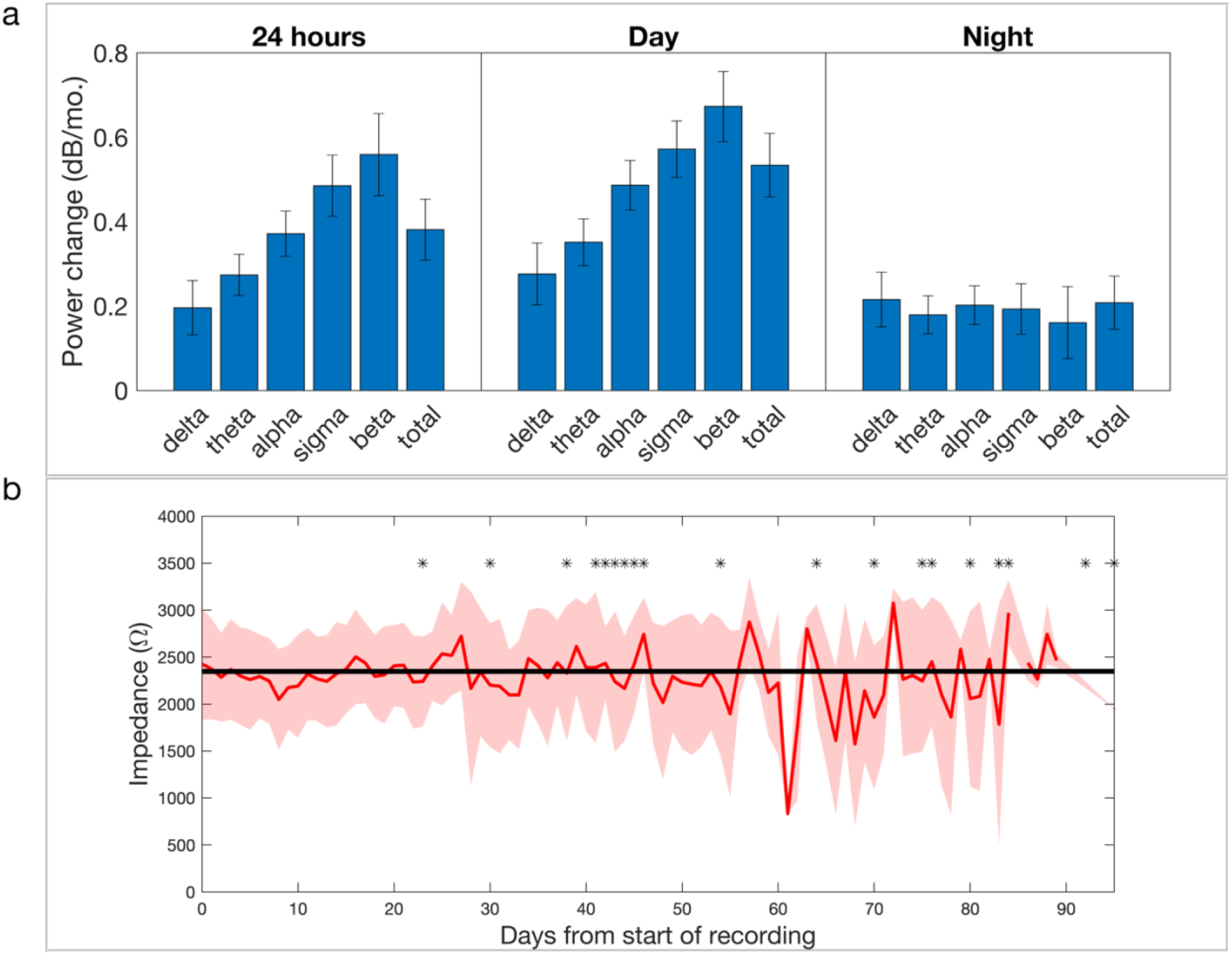
a) Results of group-level linear mixed effects (LME) models on median daily absolute powers, calculated for the 24-hour period, diurnal and nocturnal periods (left to right panels). Bars represent the coefficient of the fixed effect of time (in dB/month and kΩ/month), and error bars represent 95% confidence intervals. All were statistically significant (i.e. 95% CI not including zero). b) impedance change throughout days of the study. Red line and shaded area represent the mean and standard deviation of daily impedance measurements, respectively. Black asterisks denote the end of the recording for each subject. Black line represents the result of the group-level LME model, with an intercept of 2.35 kOhms and slope of −0.018 kOhms/month. Note that only the first 95 days of recording are represented in the graph, as only one subject recorded longer than this period, and note higher fluctuations at the end of the graph, due to fewer subjects contributing to the model.

Group-level linear mixed effects models for each frequency band power showed a small but significant increase in power with time (Figure 3a). Accounting for the full 24-hour-period, broadband power increased by 0.38 dB/month (95% CI 0.31-0.45). Power increase was larger during the day than during the night (0.53 [0.46-0.61] vs. 0.21 [0.14-0.27] dB/month, respectively). Frequency-band-specific power models showed larger positive slopes for higher frequencies than for lower frequencies for the diurnal period but not the nocturnal period. There was no significant effect of patient status on the gradient broadband power or frequency band power change (when adding the patient-time interaction term).

Electrode impedance measurements taken throughout the study, for every subject. All remained low (all below 5kOhm) and were highly stable within-subjects, with linear regression models showing long-term trends close to zero (Supplementary Figure 4). Group-level analysis showed a very small decreasing trend with time (−0.018 kOhms/month [95% CI −0.024 to −0.012]) (Figure 3b).

### Between-group comparisons of ultra long-term frequency band powers

We explored differences in frequency band power from ultra long-term recordings between healthy subjects and patients with epilepsy. No significant differences were seen when comparing median relative powers in any frequency range, between both groups (Table 1).

## Discussion

We report the largest cohort of human ultra long-term (multiple months) subcutaneous EEG monitoring, pooling over three years of recordings in total, in healthy subjects and patients with epilepsy. Signal quality metrics such as impedance and frequency-band specific power remained relatively stable over weeks and up to seven months.

We show that the sqEEG power spectra follow similar patterns to scalp EEG, with an approximate 1/f power distribution and physiological peaks present as expected. This decreasing power with higher frequencies, is similar to what is seen in scalp EEG resting state.^20,21^ In our study, we are averaging over long periods of time, encompassing activities of everyday life, including active movements and meals, which likely add myogenic artifact. This probably explains the flattening of the 1/f curve during the day in several subjects, with enhanced muscle activity, which is reflected as a broadband increase in power. Besides the different shape of the power spectrum curve, in most subjects there were clear differences in the power spectrum distribution between presumed daytime and nighttime periods. This highlights the ability of such systems to differentiate awake from sleep states.^22^

Even with long-term averaging, peaks in oscillatory activity were seen. The most prevalent diurnal peak appeared in the alpha range (e.g. HC12) and, in some patients with epilepsy, in the low alpha / upper theta range (e.g. H, E, A). We strongly suspect this corresponds to the posterior dominant rhythm, or alpha rhythm.^23^ Enhanced alpha on eye closure has previously been demonstrated with sqEEG,^13^ even with limited spatial sampling, and it is known that the alpha rhythm has a wide spatial distribution.^23^ The peak alpha frequency has been shown in numerous previous studies to be lower in patients with epilepsy^.24–26^ possibly explained by the effects of some antiseizure medications^.27^ structural network changes^25^ or, related to this, as a marker of poor seizure control^.24^ Although no significant difference in relative alpha power was seen between patients and controls in this cohort, we compared across the full 24-hour period, and have not quantified the peak alpha frequency or its variability, calling for future studies in this topic. Nocturnal power spectra showed, in the majority of subjects, a characteristic peak in the sigma range. Sigma is the frequency correlate of sleep spindles.^28^ It is interesting to note that sigma peaks appear clearer in subjects with a vertical or oblique implantation, which likely captures more midline cerebral activity where sleep spindles often predominate in amplitude. Interestingly, a sigma peak is seen in the second (more vertical) implantation of S01 (S01_2), but not in the first, horizontal / temporal implantation. Variation in sigma activity could correlate with sleep staging, and this finding may be related to the reasonable accuracy of a sqEEG sleep classifier.^22^

We found overall high stability of sqEEG over the study period for each subject. Signal stability has been investigated in implants for long-term intracranial EEG monitoring. Nurse et al. analysed subdural electrocorticography from the NeuroVista device, showing robust spectral features at group level, despite high interindividual variability.^29^ Ung et al. found, in the same dataset, a significant decline in several EEG features relevant for seizure detection and prediction (mean EEG signal powers, mean line length, energy and half-wave) at a group level in the first 100 days recording, followed by stabilization.^30^ Sillay et al. investigated ECoG electrode impedances of the NeuroPace RNS device, showing short-term (12-weeks) impedance increases, followed by stabilization in the long-term^.31^ To our knowledge, this is the first report of long-term signal stability of a minimally-invasive EEG device. We found high consistency in sqEEG electrode impedance measurements for all subjects across their periods of monitoring, from the start of the recording. This represents a significant technological advance in EEG monitoring. The low signal stability of scalp electrodes is an obvious limitation in long-term EEG monitoring, requiring regular attention by EEG technologists.

We also found overall high consistency in frequency band power throughout the recording periods, albeit with some interindividual variability. A proportion of patients showed a mild power increase with time, both across the whole spectrum and within specific frequency bands, namely more at higher frequencies during the day (Figure 3, Supplementary Figure 2). Several possible explanations could account for this effect. Even though recordings started 2-3 weeks after implantation, remodeling of tissue with changes at the electrode-tissue interface could still be seen after this period, affecting power especially at the early stage. However, examination of the shape of the long-term trends in power does not seem to show an initial increase in power, followed by stabilization, in most patients. Secondly, subjects’ behavior might change throughout the time of the study, with subjects becoming more active after having recovered from the implantation procedure.

Thirdly, long-term cyclical changes in brain activity have been described; in particular, spike rates and seizure rates have been shown to vary both at circadian but also at multi-day timescales ^32–34^. This could also be reflected in frequency band activity, or indirectly in patients’ behavior. Longer recordings might help to explore this phenomenon further. There is also the possibility for seasonal factors affecting the EEG.^35–38^

Overall, the consistency of sqEEG recordings highlights their potential usefulness not only for epilepsy, but for other applications. For example, stable signal quality makes this device useful for BCI applications. Rhythms in the beta and mu range are detectable and stable, which may be useful for sensorimotor rhythm paradigms.^8^ Inter-session variability would not be an issue anymore, nor the need for calibration data at the beginning of each session. This would significantly improve subject training time as well^.8^

We found no significant differences in median relative frequency band powers between healthy subjects and patients with epilepsy. Previous studies have shown some consistent and other inconsistent differences in EEG spectral features^,39–41^ however these studies assessed resting-state EEG, while our study assessed a grand average encompassing the full 24-hour period.

Several limitations in our study should be mentioned. There were different implant locations between healthy subjects and controls, and the number of subjects was rather low limiting group comparisons. We also restricted the consistency and between-groups analysis to consensus-defined frequency bands. Only one study subject was recorded for more than 6 months, and longer periods of monitoring will be necessary to explore the longer term stability of sqEEG.

In conclusion, overall, our findings add confidence to the reliability of sqEEG, and reinforce its potential utility in a variety of clinical and research applications. In particular, the high stationarity and minimal settling time of sqEEG shown are strong indicators that this system is well suited to chronic implantation for seizure detection and seizure forecasting.

## Data Availability

The data is currently unavailable; is planned to become available at a future date.

## Acknowledgments

This work was supported by the Epilepsy Foundation’s Epilepsy Innovation Institute My Seizure Gauge Project. MPR is supported by the NIHR Biomedical Research Centre; the MRC Centre for Neurodevelopmental Disorders (MR/ N026063/1); the RADAR-CNS project funded by the European Commission (www.radar-cns.org, grant agreement 115902). We would like to thank the Neurosurgical team (Mr. Harishchandra Srinivasan, Mr. Harutomo Hasegawa and Mr. Richard Selway) involved in the implantation procedures at King’s College Hospital NHS Foundation Trust. We would like to thank Andrea Biondi for his support with data collection at King’s.

## Disclosure of Conflicts of Interest

JDH and LSR are employees of UNEEG medical A/S. ESN and DF are employees and shareholders of Seer Medical. BHB has equity in Cadence Neurosciences and has received research devices from Medtronic Inc. at no cost. MPR has been a member of ad-hoc advisory boards for UNEEG medical A/S. PFV received a payment from UNEEG medical A/S for data annotation in an unrelated research study. No other authors have conflicts to declare.

## Ethical Publication Statement

We confirm that we have read the Journal’s position on issues involved in ethical publication and affirm that this report is consistent with those guidelines.

## Notes

### Clinical Trial

NCT04061707

### Funding Statement

This work was supported by the Epilepsy Foundation Epilepsy Innovation Institute My Seizure Gauge Project. MPR is supported by the NIHR Biomedical Research Centre; the MRC Centre for Neurodevelopmental Disorders (MR/ N026063/1); the RADAR-CNS project funded by the European Commission (www.radar-cns.org, grant agreement 115902).

### Author Declarations

London-Bromley REC (19/LO/0354) Committee of science ethics for Region Zealand (SJ‐551)

## References

1. Cook MJ, O’Brien TJ, Berkovic SF, Murphy M, Morokoff A, Fabinyi G, et al. Prediction of seizure likelihood with a long-term, implanted seizure advisory system in patients with drug-resistant epilepsy: a first-in-man study. Lancet Neurol. 2013; 12(6):563–71.

2. Duun-Henriksen J, Baud M, Richardson MP, Cook M, Kouvas G, Heasman JM, et al. A new era in electroencephalographic monitoring? Subscalp devices for ultra-long-term recordings. Epilepsia. 2020; 61(9):1805–17.

3. Sun FT, Morrell MJ. The RNS System: responsive cortical stimulation for the treatment of refractory partial epilepsy. Expert Rev Med Devices. 2014; 11(6):563– 72.

4. Kremen V, Brinkmann BH, Kim I, Guragain H, Nasseri M, Magee AL, et al. Integrating Brain Implants With Local and Distributed Computing Devices: A Next Generation Epilepsy Management System. IEEE J Transl Eng Health Med. 2018; 6:2500112.

5. Weisdorf S, Duun-Henriksen J, Kjeldsen MJ, Poulsen FR, Gangstad SW, Kjaer TW. Ultra-long-term subcutaneous home monitoring of epilepsy-490 days of EEG from nine patients. Epilepsia. 2019; 60(11):2204–14.

6. Skarpaas TL, Jarosiewicz B, Morrell MJ. Brain-responsive neurostimulation for epilepsy (RNS® System). Epilepsy Res. 2019; 153:68–70.

7. Martins IP, Westerfield M, Lopes M, Maruta C, Gil-da-Costa R. Brain state monitoring for the future prediction of migraine attacks. Cephalalgia. 2020; 40(3):255–65.

8. Abiri R, Borhani S, Sellers EW, Jiang Y, Zhao X. A comprehensive review of EEG-based brain–computer interface paradigms. J Neural Eng. 2019; 16(1):011001.

9. Di Flumeri G, Aricò P, Borghini G, Sciaraffa N, Di Florio A, Babiloni F. The Dry Revolution: Evaluation of Three Different EEG Dry Electrode Types in Terms of Signal Spectral Features, Mental States Classification and Usability. Sensors. 2019; 19(6).

10. Gu Y, Cleeren E, Dan J, Claes K, Van Paesschen W, Van Huffel S, et al. Comparison between scalp EEG and behind-the-ear EEG for development of a wearable seizure detection system for patients with focal epilepsy. Sensors (Basel) 2017; 18(1).

11. Zibrandtsen IC, Kidmose P, Christensen CB, Kjaer TW. Ear-EEG detects ictal and interictal abnormalities in focal and generalized epilepsy – A comparison with scalp EEG monitoring. Clin Neurophysiol. 2017; 128(12):2454–61.

12. Weisdorf S, Gangstad SW, Duun-Henriksen J, Mosholt KSS, Kjær TW. High similarity between EEG from subcutaneous and proximate scalp electrodes in patients with temporal lobe epilepsy. J Neurophysiol. 2018; 120(3):1451–60.

13. Duun-Henriksen J, Kjaer TW, Looney D, Atkins MD, Sørensen JA, Rose M, et al. EEG Signal Quality of a Subcutaneous Recording System Compared to Standard Surface Electrodes. Journal of Sensors; 2015.

14. Nasseri M, Nurse E, Glasstetter M, Böttcher S, Gregg NM, Laks Nandakumar A, et al. Signal quality and patient experience with wearable devices for epilepsy management. Epilepsia. 2020; 61 Suppl 1:S25–35.

15. Proix T, Truccolo W, Leguia MG, Tcheng TK, King-Stephens D, Rao VR, et al. Forecasting seizure risk in adults with focal epilepsy: a development and validation study. Lancet Neurol. 2021; 20(2):127–35.

16. Maturana MI, Meisel C, Dell K, Karoly PJ, D’Souza W, Grayden DB, et al. Critical slowing down as a biomarker for seizure susceptibility. Nat Commun. 2020; 11(1):2172.

17. Kondacs A, Szabó M. Long-term intra-individual variability of the background EEG in normals. Clin Neurophysiol. 1999; 110(10):1708–16.

18. Tenke CE, Kayser J, Alvarenga JE, Abraham KS, Warner V, Talati A, et al. Temporal stability of posterior EEG alpha over twelve years. Clin Neurophysiol. 2018; 129(7):1410–7.

19. Babiloni C, Barry RJ, Başar E, Blinowska KJ, Cichocki A, Drinkenburg WH, et al. International Federation of Clinical Neurophysiology (IFCN)--EEG research workgroup: Recommendations on frequency and topographic analysis of resting state EEG rhythms. Part 1: Applications in clinical research studies. Clin Neurophysiol. 2020; 131(1):285–307.

20. Donoghue T, Haller M, Peterson EJ, et al. Parameterizing neural power spectra into periodic and aperiodic components. Nat. Neurosci. 2020;23(12):1655–1665.

21. Voytek B, Kramer MA, Case J, Lepage KQ, Tempesta ZR, Knight RT, et al. Age-Related Changes in 1/f Neural Electrophysiological Noise. J Neurosci. 2015; 35(38):13257–65.

22. Gangstad SW, Mikkelsen KB, Kidmose P, Tabar YR, Weisdorf S, Lauritzen MH, et al. Automatic sleep stage classification based on subcutaneous EEG in patients with epilepsy. Biomed Eng Online. 2019; 18(1):106.

23. Schomer DL, da Silva FL. Niedermeyer’s Electroencephalography: Basic Principles, Clinical Applications, and Related Fields. Lippincott Williams & Wilkins; 2012. 1296 p.

24. Abela E, Pawley AD, Tangwiriyasakul C, Yaakub SN, Chowdhury FA, Elwes RDC, et al. Slower alpha rhythm associates with poorer seizure control in epilepsy. Ann Clin Transl Neurol. 2019; 6(2):333–43.

25. Yaakub SN, Tangwiriyasakul C, Abela E, Koutroumanidis M, Elwes RDC, Barker GJ, et al. Heritability of alpha and sensorimotor network changes in temporal lobe epilepsy. Ann Clin Transl Neurol. 2020; 7(5):667–76.

26. Larsson PG, Kostov H. Lower frequency variability in the alpha activity in EEG among patients with epilepsy. Clin Neurophysiol. 2005; 116(11):2701–6.

27. Ahn S-J, Kim T-J, Cha KS, Jun J-S, Byun J-I, Shin Y-W, et al. Effects of perampanel on cognition and quantitative electroencephalography in patients with epilepsy [Internet]. Vol. 115, Epilepsy & Behavior. 2021. p. 107514.

28. Goldschmied JR, Lacourse K, Maislin G, Delfrate J, Gehrman P, Pack FM, Staley B, Pack AI, Younes M, Kuna ST, Warby SC. Spindles are highly heritable as identified by different spindle detectors. Sleep. 2020; Nov 9

29. Nurse ES, John SE, Freestone DR, Oxley TJ, Ung H, Berkovic SF, et al. Consistency of Long-Term Subdural Electrocorticography in Humans. IEEE Trans Biomed Eng. 2018; 65(2):344–52.

30. Ung H, Baldassano SN, Bink H, Krieger AM, Williams S, Vitale F, et al. Intracranial EEG fluctuates over months after implanting electrodes in human brain. J Neural Eng. 2017; 14(5):056011.

31. Sillay KA, Rutecki P, Cicora K, Worrell G, Drazkowski J, Shih JJ, et al. Long-term measurement of impedance in chronically implanted depth and subdural electrodes during responsive neurostimulation in humans. Brain Stimul. 2013; 6(5):718–26.

32. Karoly PJ, Freestone DR, Boston R, Grayden DB, Himes D, Leyde K, et al. Interictal spikes and epileptic seizures: their relationship and underlying rhythmicity. Brain. 2016; 139(Pt 4):1066–78.

33. Baud MO, Kleen JK, Mirro EA, Andrechak JC, King-Stephens D, Chang EF, et al. Multi-day rhythms modulate seizure risk in epilepsy. Nat Commun. 2018; 9(1):88.

34. Viana PF, Duun-Henriksen J, Glasstëter M, Dümpelmann M, Nurse ES, Martins IP, Dumanis SB, Schulze-Bonhage A, Freestone DR, Brinkmann BH, Richardson MP. 230 days of ultra long-term subcutaneous EEG: seizure cycle analysis and comparison to patient diary. Ann Clin Transl Neurol. 2021 Jan;8(1):288–293.

35. Barbato G, Cirace F, Monteforte E, Costanzo A. Seasonal variation of spontaneous blink rate and beta EEG activity. Psychiatry Res. 2018; 270:126–33.

36. Peterson CK, Harmon-Jones E. Circadian and seasonal variability of resting frontal EEG asymmetry. Biol Psychol. 2009; 80(3):315–20.

37. Polich J, Geisler MW. P300 seasonal variation. Biol Psychol. 1991; 32(2-3):173–9.

38. Danesi MA. Seasonal variations in the incidence of photoparoxysmal response to stimulation among photosensitive epileptic patients: evidence from repeated EEG recordings. J Neurol Neurosurg Psychiatry. 1988; 51(6):875–7.

39. Drake ME, Padamadan H, Newell SA. Interictal quantitative EEG in epilepsy. Seizure. 1998; 7(1):39–42.

40. Adebimpe A, Aarabi A, Bourel-Ponchel E, Mahmoudzadeh M, Wallois F. EEG resting state analysis of cortical sources in patients with benign epilepsy with centrotemporal spikes. Neuroimage Clin. 2015; 9:275–82.

41. Pellegrino G, Tombini M, Curcio G, Campana C, Di Pino G, Assenza G, et al. Slow Activity in Focal Epilepsy During Sleep and Wakefulness. Clin EEG Neurosci. 2017; 48(3):200–8.

