## Supplementary Table 1 for "Signal quality and power spectrum analysis of remote ultra long-term subcutaneous EEG"

**Supplementary Material**

Supplementary Table 1. Baseline / Recording characteristics of study subjects.

| Subject ID | Implant Location | Recording duration (days) | Data capture rate (%) | Data capture rate - day (%) | Data capture rate - night (%) |
|---|---|---|---|---|---|
| A | LT | 30 | 86 | 95 | 93 |
| B | LT | 83 | 75 | 80 | 67 |
| C | RT | 92 | 25 | 28 | 19 |
| D | RFT | 80 | 66 | 63 | 75 |
| E | LT | 76 | 62 | 71 | 56 |
| F | LT | 84 | 91 | 87 | 94 |
| G | LT | 70 | 90 | 90 | 97 |
| H | LT | 75 | 75 | 72 | 77 |
| I | LT | 95 | 70 | 73 | 71 |
| S01 | LT | 231 | 84 | 85 | 88 |
| S01_2 | LFT | 96 | 88 | 89 | 92 |
| S02 | RT | 46 | 73 | 71 | 81 |
| S04 | RFT | 104 | 55 | 29 | 57 |
| S05 | RFT | 64 | 93 | 90 | 92 |
| S06 | LFT | 54 | 89 | 92 | 91 |
| HC_1 | RC | 43 | 75 | 75 | 74 |
| HC_2 | RC | 43 | 87 | 89 | 90 |
| HC_3 | RC | 44 | 80 | 71 | 87 |
| HC_4 | RC | 42 | 50 | 76 | 3 |
| HC_5 | RC | 45 | 87 | 85 | 94 |
| HC_6 | RC | 41 | 71 | 73 | 65 |
| HC_7 | RC | 42 | 66 | 68 | 73 |
| HC_8 | RC | 42 | 61 | 55 | 63 |
| HC_9 | RC | 38 | 52 | 54 | 49 |
| HC_10 | RC | 23 | 77 | 79 | 78 |
| HC_11 | RC | 42 | 31 | 38 | 31 |

| | | | | | |
|---|---|---|---|---|---|
| HC_12 | RC | 43 | 25 | 29 | 24 |

LT: Left temporal. LFT: left frontotemporal, RC: Right central, RT: right temporal, RFT: right frontotemporal

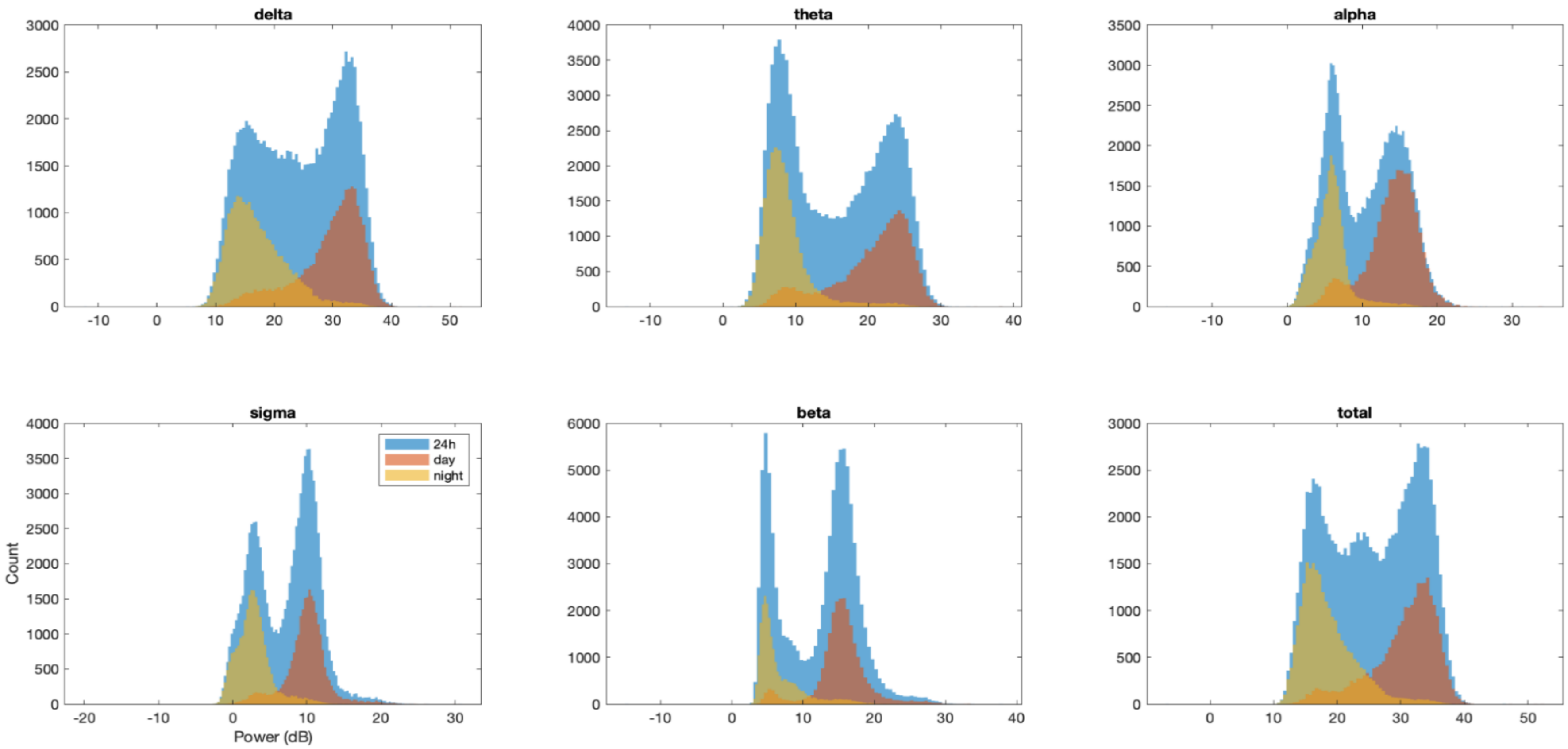


Supplementary Figure 1. Histograms of the distribution of the median absolute power values at each minute segment of the study, for each frequency band, divided into the full 24-hour period (blue), day-time (brown) and night-time (yellow) periods (example Subject I). A bimodal distribution is shown with the peaks being mostly represented by the daytime and nighttime periods.

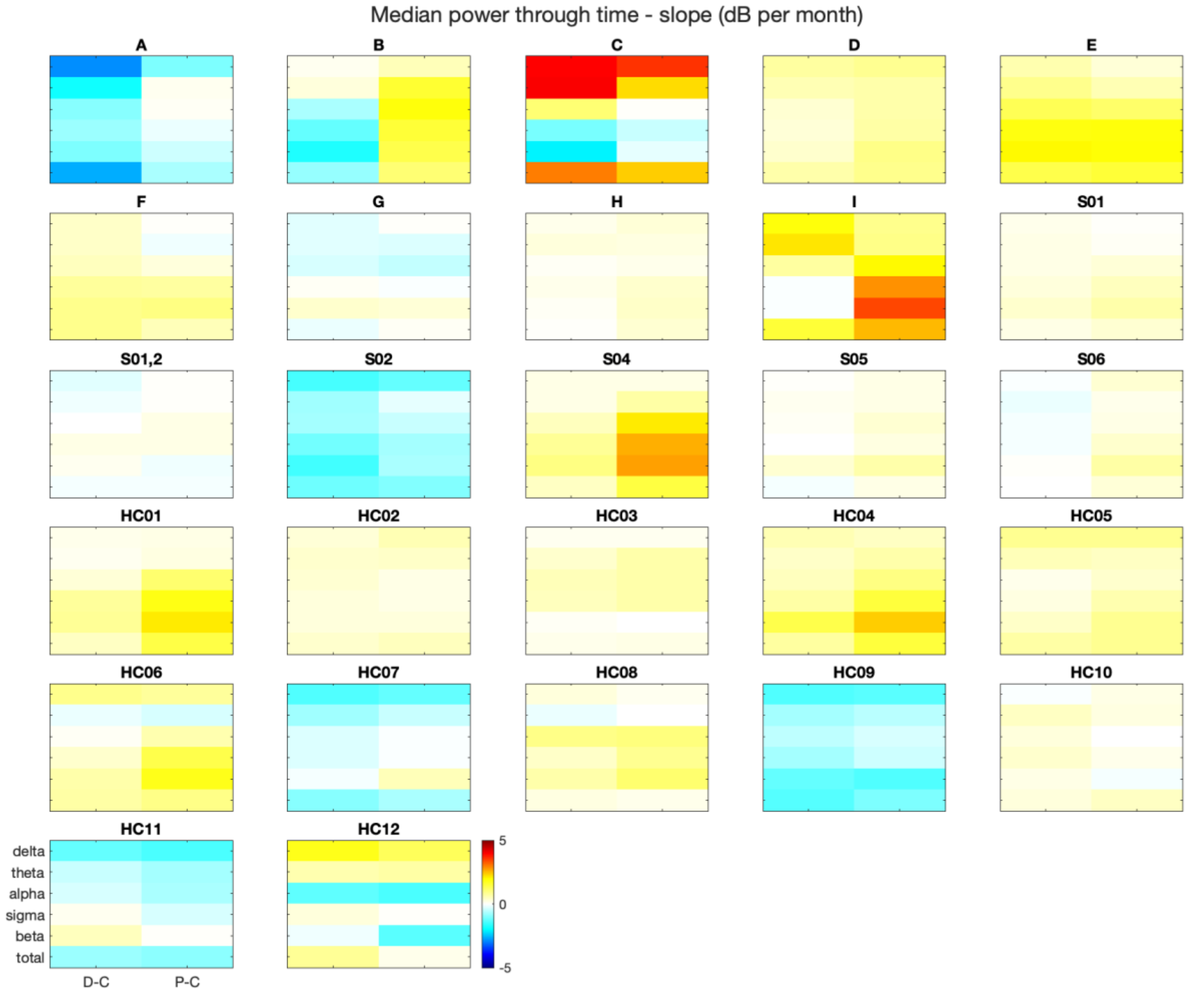


Supplementary Figure 2. Slopes (in dB/month) of individual linear regression models that fit the median daily absolute power in different frequency bands, channels and subjects.

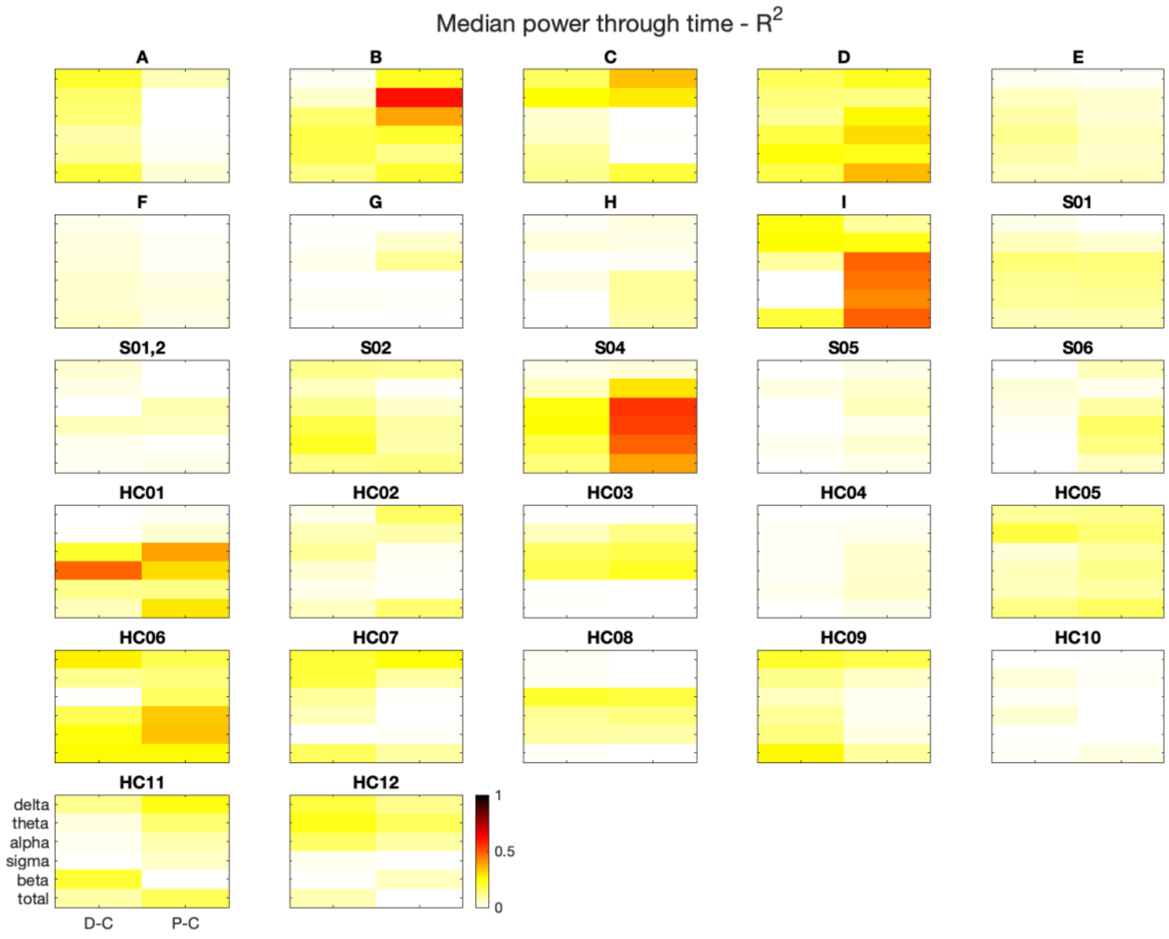


Supplementary Figure 3. $R^2$ values of the individual linear regression models (same models as in Supplementary Figure 2).

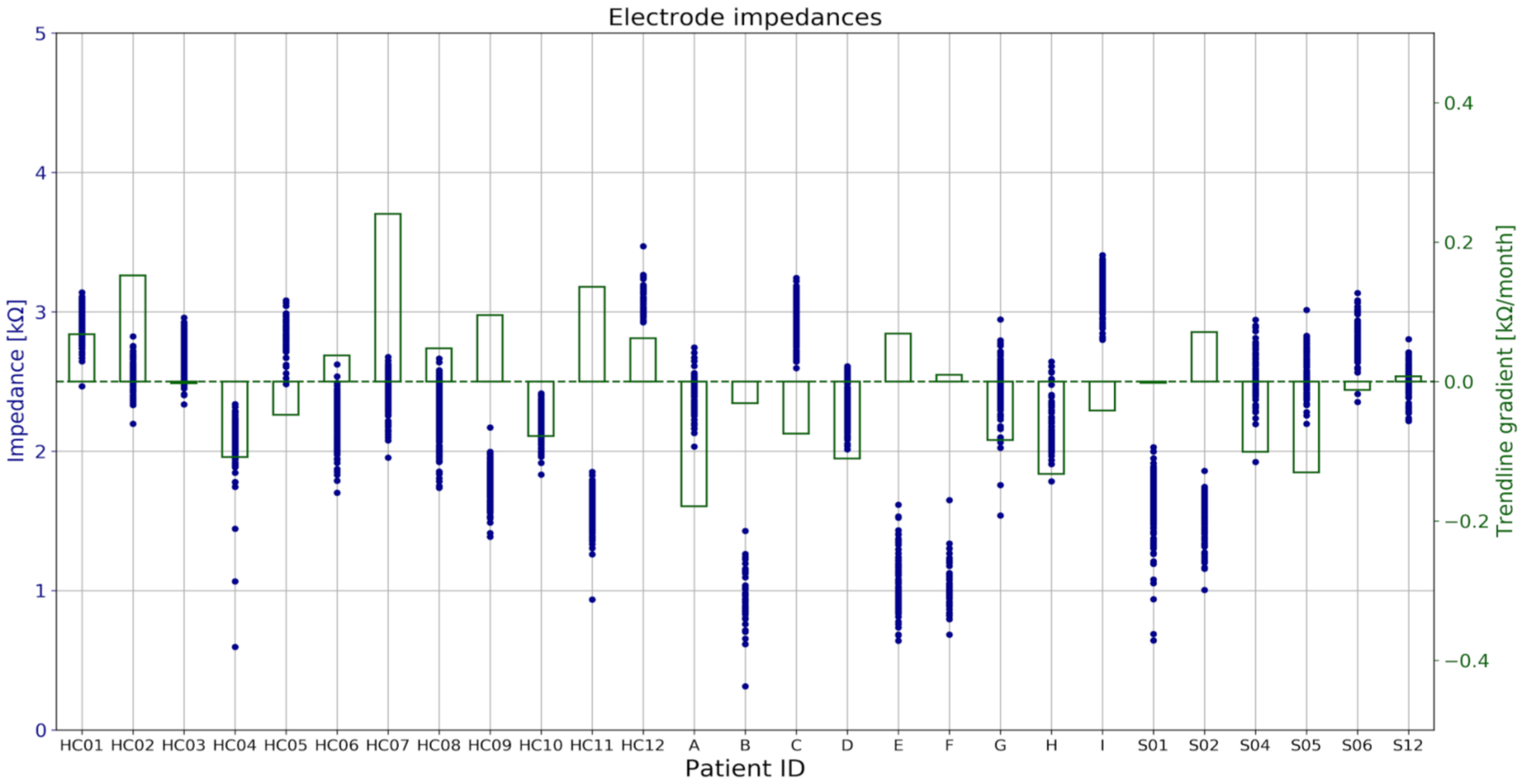


Supplementary Figure 4. The figure shows all the estimated electrode impedances (blue dots) for every patient. The gradients of patient-specific linear regression models that fit impedance changes with time are depicted in the barplots (green).
